# Adrenaline rush in athletes: Visualizing glucose fluctuations during high-intensity races

**DOI:** 10.1101/2023.05.12.23289815

**Authors:** Taira Kajisa, Toshiyuki Sakai

## Abstract

Under stressful or exciting conditions, athletes can perform beyond their typical capabilities during a so-called “adrenaline rush.” In the preliminary study by one sub-elite runner, we found that even in the fasted state, hyperglycemia occurs during high loaded running by the fact that both blood glucose and interstitial fluid glucose levels rose rapidly to 11-12 mM. This suggests that glycogen in the liver is degraded by anti-stress hormones, leading to an increase in glucose concentration. In the next, we analyzed the temporal changes in interstitial glucose concentration before, during, and after races using continuous glucose monitoring (CGM) data obtained from a total of 36 elite long-distance athletes including walking race (non-fasting state). We found that even healthy subjects recorded high glucose levels (mean 8.3 ± 1.5 mM) before the start of the race and the glucose fluctuations during the race were also recorded at 11.2 ± 2.2 mM, suggesting not only blood glucose level fluctuation due to supplementation before the races, but also due to the effects of stress hormones such as epinephrine, cortisol and glucagon. Furthermore, the mean glucose level during the daytime for the three days before the race event was significantly different by 0.3 mM (p<0.001) compared to the mean during the daytime for the three days after the race. These results suggested that efficient utilization of liver glycogen is important to keep high performance throughout the race, since the liver glycogen also consumed under stress.

## INTRODUCTION

Athletes depend on both their physical and mental conditioning to perform at the top of their ability during crucial sporting moments. Mentally, the athlete’s ability to regulate their mental strain or excitement can drastically impact the quality of their performance,^1^ while physically, the athlete must be able to expend glucose without energy depletion during long periods of competition. Excessive mental strain can cause muscle tension, which impairs performance. Conversely, excessive excitement can sometimes trigger an “adrenaline rush”^2-5^, where an athlete’s performance actually exceeds their typical ability. During competition, the muscles of high-level medium to long-distance athletes are constantly supplied with glucose for ATP production via glycolysis and the TCA cycle^6^. The athlete’s blood glucose levels can be increased by two mechanisms: (1) oral intake of sugars; or (2) hyperglycemia induced by stress. In glucose concentration, the diagnostic criteria for diabetes include a fasting blood glucose level of higher than 7 mM, a 2-h post load plasma glucose higher than 11 mM after a 75g oral glucose tolerance test (OGTT) test, or a random blood glucose ≥ 11.1 mM (Use of glycated hemoglobin (HbA1c) in the diagnosis of diabetes mellitus)^7-9^. In normal subjects, while it has been reported that athletes rarely showed hyperglycemia (>10 mM) during exercise, insulin secretion functions normally and rarely exceed 10 mM even during postprandial hyperglycemia. However, the research from bungee jumping experiments has shown that acute stress from strain can increase blood glucose levels^10^. As an athlete’s ability to control their emotions can impact their blood glucose levels and their ability to use energy effectively, a possible mechanism for hyperglycemia under stress is the acceleration of glycogen degradation in the liver by anti-stress hormones^11-14^. Taken glucose from oral intake is stored in the liver and muscles as glycogen, on the other hand, glycogen degradation is accelerated by the secreted anti-stress hormones due to excitement and tension leading to utilizing ATP production from muscle glycogen and releasing glucose in blood from liver glycogen^15-20^. Thus, it is required for athletes to control their emotions due to tension and excitement before the real race and to use energy effectively.

It is important for athletes to monitor their blood glucose levels during exercise to control energy balance and maintain performance^21-23^. Recent studies have examined the impact of continuous glucose monitoring (CGM) system on athletic performance^24-29^. In one study involving a sub-elite British soccer team, 70% players performed in a hyperglycemic state of ≥ 6 mM with interstitial glucose level elevations during matches up to 10 mM^27^. Suzuki et al. also reported that athletes can experience interstitial glucose concentration increases up to 10 mM during long-distance running^28^. These studies are limited because they included little information regarding the balance of nutrient intake and energy expenditure or the impact of an athlete’s mental status on blood glucose concentration. Additionally, with the goal of this study being to use CGM data during long-distance races in college track and field athletes, these studies had small sample sizes and did not include high-level athletes. We aimed, by monitoring glucose levels in the body before, during, and after highly competitive—and potentially mentally stressful—races, to examine the energy consumption of top runners.

## METHODS

### Study subjects and collection of CGM data

In this study, interstitial glucose levels in all runners were continuously monitored using FreeStyle Libre^®^ flash glucose monitoring systems (FreeStyle Libre system; Abbott Diabetes Care, Alameda, CA) which apply patch-type CGM sensor under the skin of the upper arm. CGM data were collected for a total of 37 study subjects (one sub-elite subject for preliminary study, and for the main study, the total of 36 elite runners belonging to track and field in Toyo University).

To prove the correlation between blood glucose and interstitial fluid glucose levels during intense running and evaluate the measurement error and temporal discrepancies in blood glucose levels and interstitial fluid glucose concentrations by CGM during exercise, one healthy sub-elite runner (male, early 40’s, HbA1c: 5.1, fasted plasma glucose level: 5.4 mM) measured blood glucose levels by Glutest Neo Alpha^®^, which is an enzymatic glucose sensor with finger prick using a Gentlet^®^ (Sanwa Kagaku Kenkyusho Co. Ltd., Nagoya, Japan)) in parallel with the CGM measurement as a preliminary test. We examined the blood glucose levels during a race for the sub-elite subject who fasted for 12 h from 19:00 the previous day ran for 5 km distance equipped with CGM and with measured blood glucose level using self-monitroing of blood glucose (SMBG) every several tens of minutes. This athlete completed an OGTT after finishing his high-intensity running. Subsequently, for long-term monitoring of glucose levels before and after a full marathon race, this sub-elite completed a full marathon race during that period while wearing the CGM for 18 days.

In the main experiment, 20 college elite athletes (male, early 20’s) who specialize in long-distance running or race walking participated in track, road race, and relay competitions while equipped with CGM, and a total of 36 CGM data were collected through the serious races without the fasted state. Participants were recruited from five months prior to each race. They satisfied the entry criteria for intercollegiate competitions as well as top long-distance races in Japan. Furthermore, the selection was made from athletes who had been chosen as the top performers within their respective teams. Detail regarding the athletic events included in this study are shown in Table 1.

**Table 1.** Collected data information on athletes in actual races and their type of competition.

| Sample number | Type of competition | Race distance (km) |
| --- | --- | --- |
| n = 2 | Run, track | 10 |
| n = 6 | Run, road (relay) | 5.8, 6.2, 6.4, 8.0, 8.5, 10.2 |
| n = 8 | Run, road (relay) | 9.5, 11.1, 11.8, 11.9, 12.4, 12.8, 17.6, 19.7 |
| n = 8 | Run, road (relay) | 20.8, 20.9, 21.3, 21.4, 23.0, 23.1 |
| n = 1 | Run, cross country | 10 |
| n = 2 | Race walk, road | 20, 35 |
| n = 3 | Run, track | 10 |
| n = 1 | Race walk, road | 10 |
| n = 2 | Run, road | 21.0 |
| n = 3 | Run, track | 5 |
| n = 36 | 16360 plots from 3 days before the race day to 3 days after the race day |  |

All procedures were approved by the ethical review committee for medical research involving human subjects at Toyo University (TU2021-006). Data collection was carried out for all athletes after providing an explanation of the experimental methods and associated risks, and obtaining written informed consent through signed consent forms.

### Statistical analysis of CGM data

To eliminate individual differences and biases in the numerical data, the CGM plots were statistically analyzed from a sufficient amount of data across various long-distance events to ensure robust results. In measuring glucose levels, the CGM automatically plotted 15-minute averages of glucose values per minute. In sub-elite runners, glucose variability data were plotted for 18 days. 36 glucose variability data for elite runners were plotted for one week, three days before and after each match day. Plotted glucose data was analyzed by statistical methods. Statistical analyses were performed using Microsoft Excel 2016 (Microsoft Corp., Redmond, WA, USA). We compared glucose levels before, during, and after races, using a one-way analysis of variance (ANOVA). Following the results of ANOVA, we performed pairwise comparisons between timepoints using a two-tailed Welch’s *t*-test. We also calculated summary statistics for glucose levels during each timepoint (mean, median, standard deviation, interquartile range).

## RESULTS

### Estimation and comparison of glucose level measured by self monitoring of blood glucose (SMBG) and CGM during high-intensity running

In this study we found that CGM can be used to monitor blood glucose concentrations for high-level runners during races and these athletes’ glucose levels differ before-, during-, and after-races. First, we evaluated the measurement error and temporal discrepancies in blood glucose levels by SMBG and interstitial fluid glucose concentrations by CGM during exercise. Figure 2 displays the blood glucose data for a healthy sub-elite runner (male, 40 s, HbA1c: 5.1, fasted plasma glucose level: 5.4 mM) during fasting, running, and subsequent OGTT testing.

**Figure 1.** Mechanism of blood glucose elevation by secreted anti-stress hormones promoted from stress factors such as tension and excitement.

**Figure 2.** Time course graph of glucose level fluctuation by self-monitoring blood glucose (SMBG) and continuous glucose monitoring (CGM) during high-intensity running and glucose tolerance test after 12 h of fasting (Red: SMBG, Blue: CGM).

Figure 2 also shows the participant’s glucose levels over time using both SMBG and CGM. The participant ran at an average pace of 4 min/km with maximum heart rate of 175 bpm during 12 h period fasting state and subsequent oral glucose ingestion after running. During high-intensity running, the maximum glucose level recorded 12.2 mM for SMBG and 11.3 mM for CGM despite of fasting conditions. Immediately after running, when glucose levels returned to the normal level, the participant completed a glucose endurance test. After ingesting glucose, the participant’s blood glucose levels were recorded at a maximum of 7.7 mM in SMBG and 6.3 mM in CGM after approximately 30 min and returned to normal levels after 1 h. The average blood glucose value measured by CGM was 1.5 mM lower than that of SMBG. Additionally, we observed a delay of about 15 min in the speed of glucose rise during running in the CGM measurement compared to the SMBG measurement.

In the next experiment, interstitial fluid glucose monitoring data of the same male subelite athlete was collected for 18 days before and after the full marathon race with non-fasting state (Fig. 3). First, we focused on sleeping time between 1 am and 7 pm and found the CGM values were about 3.5 mM lower the day before the race. CGM values during sleeping time on race day were higher than the average values and did not fluctuate. Then, the participants’ glucose level increased sharply up to 10 mM just before the start time of the race. During the race, the participant’s CGM values were consistently above 10 mM with a maximum value of 11.7 mM during the race’s midpoint. The participant’s CGM values were consistently above 7 mM until 4 h after finishing the race. On the night after the race, he slept with extraordinarily higher blood glucose values between 5.7 and 9.6 mM. Compared with the average CGM values during the race (average pace of 4:14 min/km) and the CGM values in the full marathon practice at relatively low intensity (average pace of 6:16 min/km), it was cleared that the CGM value was quite higher during the race, while the values during the practice remained within 5 to 6 mM and the average value was 5.4 ± 1.1 mM (Fig. S1).

**Figure 3.** Interstitial glucose concentration plots of 1 sub-elite runner before and after a full marathon race (Blue: 1 day before the race, Red: race day, Green: 1 day after the race). Average, average plus a standard deviation, and average plus two standard deviations are indicated as pink, yellow, and skin color line, respectively.

### Statistical analysis of blood glucose levels in elite long-distance runners during races

We obtained interstitial glucose data using CGM device from elite runners and race walkers at various locations and times, which are 5000 and 10000 m track races, 5.8 to 35 km road races, and relay races. Table 1 shows the date and time when the sample data was collected, the number of subjects, the type and location of the race, and the distance. A total of 16360 glucose data plots were collected from 36 athletes between early 20’s. All athletes were considered healthy based on their fasting blood glucose and HbA1c levels collected during their physical examinations.

Figure 4 shows real-time data on the variation in mean body glucose levels by CGM from 2 h before the start to post-race for representative athletes in various competitive events. Although individual differences were observed in pre-race glucose variability, many athletes demonstrated a pattern of glucose elevation during the race and acute elevation of glucose levels just before starting. Many athletes also demonstrated an increase in glucose approximately 90 min before their race, which corresponded with the timing of a supplemental meal. The athletes’ mean glucose levels also continued to rise gradually during the race. After the race ended, a sharp drop in glucose levels was characteristically observed within an hour.

**Figure 4.** Changes in interstitial fluid glucose concentration from long-distance elite athletes before/during/after race competitions of different disciplines (a: 10 km running track, b: 12.8 km relay on road, c: 21.1 km half-marathon on road, d: 35 km race walk on road). The pink band indicates the running time with the start time as 0 min.

Figure 5a plots the glucose levels for each individual participant from 3 days before and after the actual races. In Fig. 5b, we analyzed glucose levels around the race by dividing CGM readings into 1 h before the race, during the race, and the 1 h after the race. The overall mean value of blood glucose was 6.1 ± 1.3 mM. The mean value of glucose rose to 8.3 ± 1.5 mM at 1 h before the race and increased to 11.2 ± 2.2 mM during the race. Mean glucose levels then dropped to 6.7 ± 1.7 mM at 1 h after the race. The median values were 8.3 mM (IQR: 1.4 mM) at 1 h before the race, 11.1 mM (IQR: 2.8 mM) during the race, and 6.6 mM (IQR: 2.7 mM) at 1 h after the race. These data indicate that glucose values were higher before and during races compared to the other timeframes. Athletes’ glucose levels before and after the race day were also compared. Figure 5c shows the distribution of daily glucose fluctuations during the 3 days before and after the race. Daily glucose levels for the 3 days before the race were 6.4 ± 0.7 mM, whereas daily glucose levels for the 3 days after the race were 6.1 ± 1.1 mM. The difference of 0.3 mM in daily glucose values for the three days before and after the race was statistically significant with a p-value of <0.001.

**Figure 5.** Statistical results and analysis from elite long-distance athletes in the real competition. (a): Actual data plots of interstitial glucose level for 7 consecutive days before and after 3 days of the race day. (b): The results in statistical analysis of interstitial glucose data before 1 h (blue), during the race (red), after 1 h (green), and total data (gray). (c): Comparison of statistical data with before 3 days (blue) and after 3 days of the race (green).

## DISCUSSION

The phenomenon of increased blood/interstitial glucose levels during exercise has been previously reported^27^. While most of these studies were conducted during controlled exercise tests, which provides little information about the potential influence of mental stress or excitement on blood glucose levels, it has been reported that increased epinephrine in the body is correlated with elevated blood glucose levels during exercise^30^. However, in this study, we used CGM to monitor body glucose levels in real-time during actual racing competitions. Therefore, at first in this report, blood glucose and interstitial fluid glucose values were compared during high-intensity running using one sub-elite runner as a preliminary test. We observed an approximately 15-minute lag for CGM values compared to SMBG values (Fig. 2). We inferred this time discrepancy reflected a displacement in the time of fluid seepage from the blood to the interstitial fluid. The correlation between blood glucose and interstitial fluid glucose concentrations and the time lag in the variation of blood glucose and interstitial fluid glucose concentrations with freestyle libre were consistent with previous reports^31,32^. Additionally, we found that both SMBG and CGM detected hyperglycemia during and after one participant’s race despite his fasted state (Fig. 2). These results suggest that the increase in blood glucose levels during high-intensity running could be due to glucose release by the breakdown of glycogen in the liver caused by the secretion of anti-stress hormones since the low potential for oral glucose source supply. While the results of this study are consistent with this mechanism of hyperglycemia, it is known that plasma concentrations of glucagon and adrenaline increase when the brain recognizes a decrease in glucose concentration, which causes hyperglycemia^33^, and, because we observed glucose concentration above 10 mM before the start of the race, this level continued to rise up to 11.7 mM during the race. We believe this increased blood glucose concentration may have been caused by an increase in anti-stress hormones due to tension and excitement associated with the competition (Fig. 3). It has been reported that moderate exercise lowers body glucose levels^28^. While, supporting the importance of oral glucose supplementation during races is the fact that healthy subjects continued to maintain a hyperglycemic state of more than 8 mM (mean +2σ) for as long as 6 h before and after the marathon race, it has additionally been suggested that elite athletes consume most glucose in the blood during competition leading to increased liver glycogen degradation.

In the next experiment, from the CGM data of the total 36 elite runners in serious race events, their glucose variability made it possible to determine their state of tension and excitement before, during, and after the race. In Figure 4, the real-time data on glucose variability were shown for races of different distances, locations, and competitions. In terms of the results, the data range from personal best records (Fig. 4a and 4c), including world record (Fig. 4d), to personal worst records (Fig. 4b), suggesting the possibility of a correlation between the results and the glucose patterns. Our sample size of 36 subjects and 16360 points were sufficient for this statistical analysis (Fig. 5a). From the evaluation of the distribution at a different stage of the serious race, the glucose elevation was observed 1 h before the race, presumably due to tension and excitement, which differed by more than 2 mM compared with the mean value of the total glucose level from 3 days before the race and 3 days after (Fig. 5c). This pre-race glucose distribution value was assumed to be due to excitement and tension, since the athletes consumed the pre-race supplementation 1.5 to 2 h before the start time of the race, and the glucose elevation due to the supplementation did not exceed 8 mM on average (Fig. 5b). Moreover, glucose values were nearly twice as high compared to normal values during the race at 11.1 mM. Usually, the test collaborators have trained at the same pace of the race one week before.

Although the pattern of glucose variability in the high-intensity practice and that of the race were similar, it was significantly higher in the race event during running (Fig. S2). Glycogen is stored up to about 100 g in the liver, whereas 500 g is stored in muscle cells^34-36^. However, it has been reported that glycogen stored in muscle cells cannot release glucose into blood vessels^15^. In other words, the results of this study suggested that the only the factors in raising the glucose level were the glycogen source in the liver, which was heavily consumed during the main race, in addition to the amount of carbohydrates by oral ingestion. In addition, the average glucose values at a daytime were higher before the race, not only one hour before the start, but also three days before, suggesting that glucose consumption was increasing through daytime. Mullin *et al*. found that the glucose consumption of 19 female heptathlon athletes was 339 g/day^37^. For this amount of glucose consumption, carbohydrates are released successively into the blood by oral and glycogen consumption to maintain a normal blood glucose level of around 5 mM. Namely, assuming that a nervous or excited runner raises his/her glucose level to about 10 mM before the race, and then to about 15 mM during the race, he/she would be releasing about 70 g of glucose into the blood from 1 h before the race to 1 h during the race. Thus, it is estimated that liver glycogen may be depleted during exercise in extreme tension and excitement. It has been reported that exercise at 75% of maximal oxygen uptake for at least 150 min does not cause hypoglycemia and continued to work for 180 min up to complete exhaustion if muscle glycogen levels are sufficient^38^, and further that people can move for nearly 90 min without hypoglycemia even when muscle glycogen levels are considerably depleted^39^. From these reports, whereas depletion of glycogen in the liver during exercise makes it difficult to maintain blood glucose levels or increases the likelihood of metabolic system switches to gluconeogenesis, which in turn increases the risk of failure, depletion of glycogen in the liver does not mean that the athletes will collapse due to hypoglycemia even if glycogen in the liver is depleted.

There exist certainly many associated factors in blood glucose levels during exercise, not limited the blood glucose factor. For example, it is also important how re-metabolized glucose levels (e.g. from lactate) are accrued during all competitive races in athletes^40,41^. However, the only biomarker that can monitor in real-time during the actual race is the interstitial fluid glucose level in current, so that it is desirable to develop continuous lactate monitoring in the future. Furthermore, continuous glucose monitoring should be combined with heart rate and maximal oxygen consumption and analyzed in correlation to reveal running events occurring in the blood. The conclusion from the continuous glucose monitoring in this research is that it is necessary to consume the limited glycogen in the liver efficiently in addition to supplying sugar through oral intake and by controlling the tension and excitement through the race in order to maintain high performance in the race.

## CONCLUSION

This study suggested that glucose level fluctuates due to the effects of stress hormones such as epinephrine, cortisol, and glucagon. To bring out the full power of the athletes’ abilities and keep their energy up during the real race, efficient utilization of liver glycogen while controlling excitement and tension is important to keep high performance throughout the race since the liver glycogen is also consumed under stress.

## Data Availability

All relevant data are within the manuscript and its Supporting Information files. All data produced are available online at 10.6084/m9.figshare.22807130.

https://doi.org/10.6084/m9.figshare.22807130.

## ACKNOWLEDGEMENTS

Part of this work was supported by HAKUJU INSTITUTE FOR HEALTH SCIENCE Co., Ltd., Nisshinbo Holdings Inc., and Japan Keirin Autorace Foundation. The authors wish to thank all of the healthy athletes who participated in the project.

## SUPPORTING INFORMATION

The Supporting Information is available free of charge at DOI:***.

### Author Information

**Corresponding Author**

**Taira Kajisa** – Graduate School of Interdisciplinary New Science, Toyo University, 2100 Kujirai, Kawagoe, Saitama 350-8585, Japan; Phone: +81-49-239-2375;

### Notes

The authors declare no competing interests.

