## Supplemental File for "Adrenaline rush in athletes: Visualizing glucose fluctuations during high-intensity races"

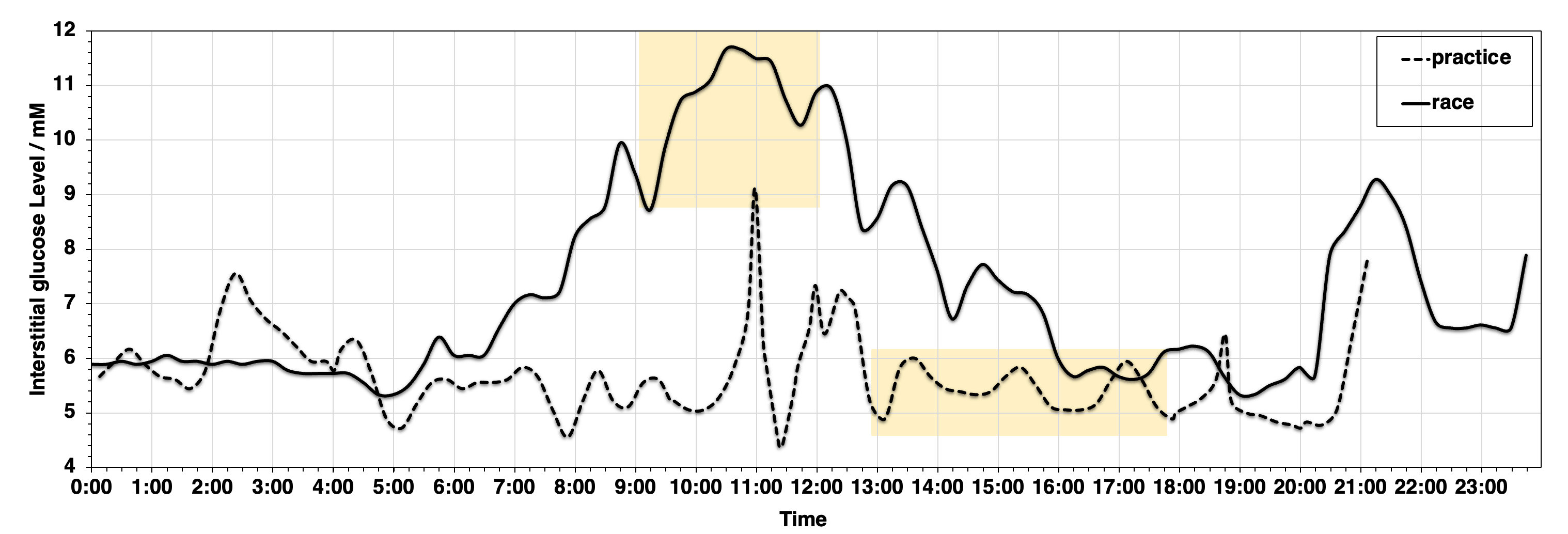

**Figure S1.** Interstitial glucose concentration curves of the sub-elite runner during a full marathon race and practice (black line: race day, dotted line: practice day). The yellow band indicates the running time of full marathon.

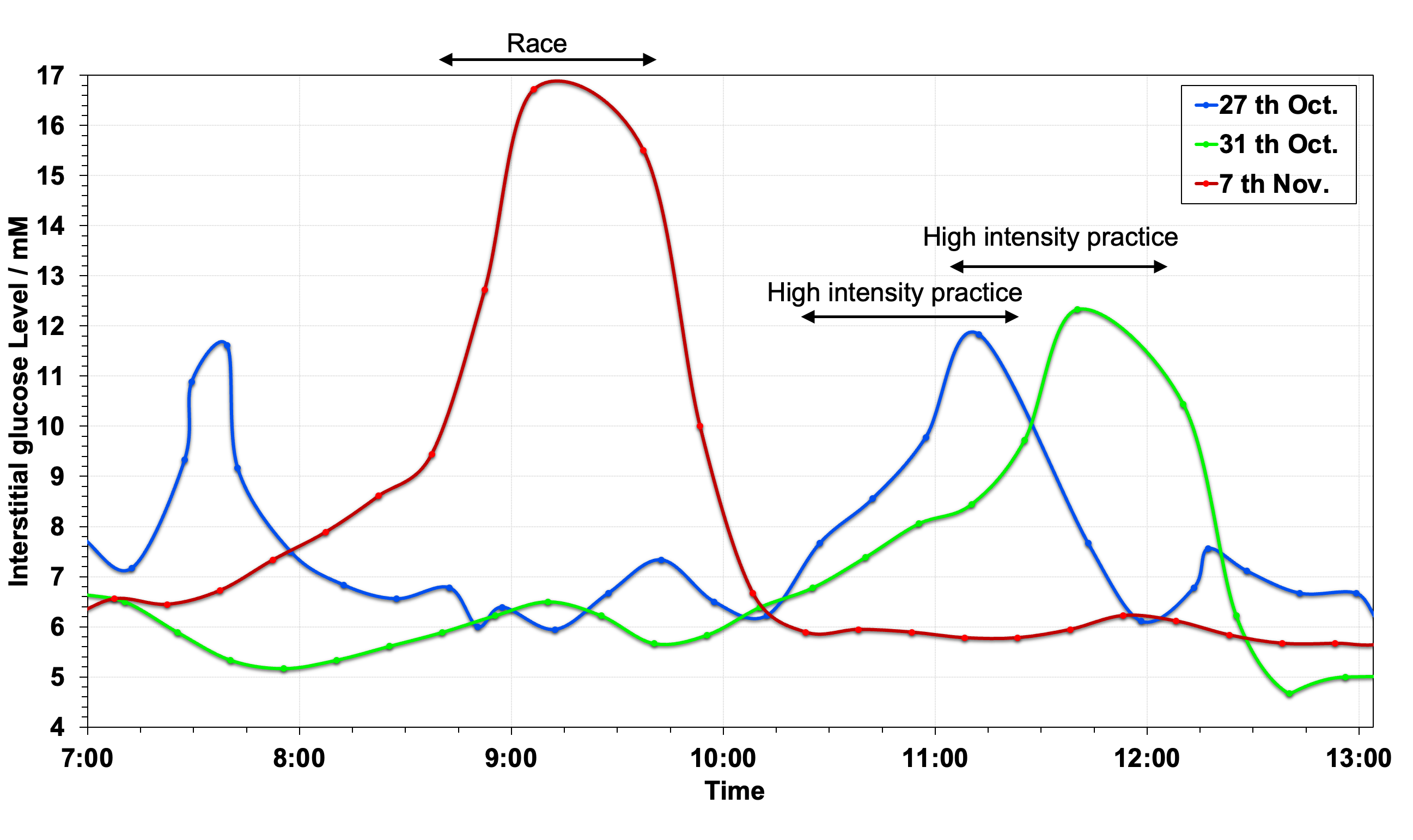

**Figure S2.** Comparison of glucose levels in a long-distance elite athlete between race and high intensity practices (Red: race, Blue and Green: high-intensity practice).
